# Differences in progression by surgical specialty: a national cohort study

**DOI:** 10.1101/2021.05.10.21256963

**Authors:** Carla Hope, Jonathan Lund, Gareth Griffiths, David J Humes

## Abstract

The aim of surgical training across the ten surgical specialties is to produce competent day one consultants. Progression through training is assessed by the Annual Review of Competency Progression (ARCP).

**Objective:** This study aimed to examine variation in ARCP outcomes within surgical training and identify differences between specialties.

**Design:** A national cohort study using data from United Kingdom Medical Education Database (UKMED) was performed. ARCP outcome was the primary outcome measure. Multi-level ordinal regression analyses were performed, with ARCP outcomes nested within trainees.

**Participants:** Higher surgical trainees (ST3-ST8) from 9 UK surgical specialties were included (vascular surgery was excluded due to insufficient data). All surgical trainees across the UK with an ARCP outcome between 2010 to 2017 were included.

**Results:** Eight thousand two hundred and twenty trainees with an ARCP outcome awarded between 2010 and 2017 were included, comprising 31,788 ARCP outcomes. There was substantial variation in the proportion of non-standard outcomes recorded across specialties with general surgery trainees having the highest proportion of non-standard outcomes (22.5%) and urology trainees the fewest 12.4%. After adjustment, general surgery trainees were 1.3 times more likely to receive a non-standard ARCP outcome compared to trainees in T&O (OR 1.33 95%CI 1.21-1.45). Urology trainees were 36% less likely to receive a non-standard outcome compared to T&O trainees (OR 0.64 95%CI 0.54-0.75). Female trainees and older age were associated with non-standard outcomes (OR 1.11 95%CI 1.02-1.22; OR 1.04 95%CI 1.03-1.05).

**Conclusion:** There is wide variation in the training outcome assessments across surgical specialties. General surgery has higher rates of non-standard outcomes compared to other surgical specialities. Across all specialities, female sex and older age were associated with non-standard outcomes.

**Article summary:** *Strengths and limitations:*

- This is the first study investigating factors affecting ARCP outcome across all surgical specialities.
- The major strength of the study is the large sample size comprising all higher surgical trainees between 2010 and 2017.
- Unlike previous studies this study uses data from reliable sources and is not dependent on survey data.
- Limitations include the inability to investigate the causes behind our findings due to the nature of the analysis.

## Introduction

Surgical training in the UK is comprised of ten specialties: cardiothoracics, ENT, general surgery, neurosurgery, oral and maxillofacial surgery (OMFS), paediatric surgery, plastic surgery, trauma and orthopaedics, urology and vascular. Successful entrants to surgical specialty training in the UK are enrolled into a six year programme within their specialty of interest, termed Higher Surgical Training (HST). This follows a minimum of four years clinical experience after medical school graduation. Specialty training is mapped to a national curriculum which defines the levels of speciality specific knowledge and skills required for completion of training around academic, procedural and clinical competence[1]. Progression to the next stage of training is dependent on meeting curriculum targets and is assessed by the Annual Review of Competency Progression (ARCP)[2]. ARCP is the formal and structured evaluation of a portfolio of evidence to ensure a doctor has achieved the required competencies to enable satisfactory progression through each stage of medical training[2]. It can be used as a marker of progression. The end point of successful completion of surgical training is the award of Certificate of Completion of Training (CCT) which allows entry onto the General Medical Council specialist register[3].

The aim of surgical training is the same across all specialties: to enable surgeons to acquire the curriculum standards and the professional responsibilities to practice as a day one consultant surgeon[4]. If the training programme is meeting the needs of trainees, we would expect similar proportions of ARCP outcomes across all surgical specialties. The GMC is committed to promoting excellence in the delivery of postgraduate medical education and therefore it is important to ensure consistency of training standards and assessment across surgical training[5].

Differential attainment within surgical training is found internationally[6-8]. Other studies have demonstrated differences in surgical training outcomes by gender, age and place of medical qualification[9], however to date there have been no studies comparing ARCP outcomes across surgical specialties.

## Aim

To examine variation in ARCP outcomes within UK surgical specialty training and identify any differences between specialties.

## Method

We performed a, cohort study using anonymised data from the UK Medical Education Database (UKMED), approval number 3506[10]. UKMED collates data on sociodemographic and educational information for UK and international medical students[11] and follows these students through their medical training, adding data as they progress. The database includes all UK medical students commencing studies in 2002 onwards, and from 2012 onwards, those graduating from non-UK medical schools and entering UK postgraduate training are included. For each graduate, ARCP outcomes are linked to demographic information. These data can be modelled to assess the differences in ARCP outcomes between surgical specialities. These data have been used to study outcomes and progression in medical specialties and foundation programme extensively and have been shown to be reliable[12-14].

### Patient and Public Involvement

Patient and public involvement was sought given the educational nature of the study and the fact no patients were included in the study.

### Outcome Measure

The outcome measure in this study was ARCP outcome. ARCP outcome was converted into an ordinal category following which has previously been validated[15].

- Group 1: “Satisfactory progress/training completed”: ARCP outcomes 1 and 6 (see Table 1)
- Group 2: “Insufficient evidence presented”: ARCP outcome 5
- Group 3: “Targeted training required (no extended time)”: ARCP outcome 2
- Group 4: “Extended training time required/left programme”: ARCP outcome 3 or 4

**Table 1.** ARCP Outcomes. Standard outcomes are ARCP outcomes 1, 6, and 7.1. Non-standard outcomes are ARCP outcome 2, 7.2, 3 and 4. Outcome 5 is awarded when insufficient evidence is presented with a window of 2 weeks for the trainee to provide missing evidence before a standard or non-standard outcome is awarded, Outcome 8 is awarded when out of programme and neither this nor any outcome 7 were included in the analysis.

| **Outcome** | **Description** |
| --- | --- |
| 1 | Satisfactory progress- achieving progress and the development of competences at the expected rate |
| 2 | Development of specific competences required- additional training time not required |
| 3 | Inadequate progress- additional training time required |
| 4 | Released from training programme- with or without specified competences |
| 5 | Incomplete evidence presented- additional training time may be |
|  | required |
| 6 | Gained all required competences- will be recommended as having completed the training programme (core of speciality) and if in a run-through training programme or higher training programme, will be recommended for award of a CCT or CESR/CEGPR |
| 7 | Fixed-term posts (split into 1-4 as above) |
| 8 | Out of Programme for clinical experience, research or a career break (OOPE/OOPR/OOPC) |

Any non-training outcomes were not included in this analysis, for example those out of programme for research or experience.

### Cohort Derivation

All trainees (ST3-ST8) from cardiothoracics, ENT, general surgery, neurosurgery, OMFS, paediatric surgery, plastic surgery, trauma and orthopaedics and urology with an ARCP outcome recorded between 2010 and 2017 were included. Trauma and orthopaedic surgery was taken as the reference category in the regression analyses as this had the largest number of outcomes. In the event of more than one ARCP outcome within 12 months, the first chronological outcome was used. It was not possible to include vascular surgery as this only became a separate specialty in 2013 and the number of outcomes was too small (N=189).

### Predictor Variable Selection

Age, sex, region of primary medical qualification, less than full time training and year of ARCP were selected as confounding factors due to the variation in these demographics across specialities. Region of primary medical qualification was categorised into UK, European Economic Area and International Medical Graduates (IMG)[16]. Less than full time training requires the same standard of competencies to be met at ARCP, however the training time may be longer.

### Statistical Analysis

Descriptive statistics were derived to describe the satisfactory and non-standard cohorts. Demographic data was rounded to ensure anonymity as per guidance from the Higher Education Statistics Agency[17]. Missing data was coded as a separate category within each variable. Chi squared test was used to compare the difference in the proportion of satisfactory outcomes compared to general surgery, as general surgery had the highest proportion of non-standard outcomes. Predicted probabilities of obtaining each ARCP outcome were calculated for each specialty to allow comparison across all specialities and displayed in a caterpillar plot using Stata post-estimation commands. A multi-level ordinal regression was performed, with a random intercept at the trainee level. This takes into account the fact that trainees had multiple ARCPs over time. Wald’s test and likelihood ratio tests were used to obtain p values.

A multi-level multiple ordinal regression was performed to investigate the odds of non-standard ARCP outcome by specialty. Odds ratios with 95% confidence intervals adjusted for sex, age, region of primary medical qualification, less than full time training and year of ARCP were reported. P values <0.05 were taken as significant. Analyses were performed using Stata version 15 (StataCorp LLC, College Station, TX, USA)[18].

## Results

The analysis included 8,220 trainees across all specialties, comprising 31,788 ARCP outcomes. Seventy six percent of all trainees were male. The proportion of female trainees varied from 15% in trauma and orthopaedics to 49% in paediatric surgery (Table 2). The median age at ARCP was 35 years (IQR 32-37). The majority of trainees (81%) had attended a UK medical school. Eighty two percent (25,991/31,788) of the outcomes were satisfactory, 8% provided insufficient evidence, 6% required targeted training and 4% either required extended training time or were asked to leave the programme (Table 3). Between 2010 and 2017, of the non-standard outcomes, 1.3% (75/5797) were Outcome 4 (released from training programme). Between 2010 and 2017 standard outcomes reduced from 85% to 78% (p <0.001).

**Table 2.** Demographics by specialty

|  | **General surgery** | **Trauma and orthopaedics** | **Plastic** | **Urology** | **Neurosurgery** | **ENT** | **Maxillofacial** | **Cardiothoracic** | **Paediatric** |
| --- | --- | --- | --- | --- | --- | --- | --- | --- | --- |
| **Age at ARCP (years, IQR)** | 35 (33-38) | 33 (30-36) | 36 (33-38) | 35 (32-37) | 34 (31-37) | 36 (33-38) | 38 (36-41) | 37 (33-40) | 35 (33-38) |
| **Sex (M:F)** | 1780:850<br>(68%) | 2130:380<br>(85%) | 435:215<br>(67%) | 535:185<br>(74%) | 415:100<br>(81%) | 295:110<br>(73%) | 240:55<br>(81%) | 225:55<br>(80%) | 115:110<br>(51%) |
| **PMQ region** |  |  |  |  |  |  |  |  |  |
| UK | 2035 (78%) | 2060 (82%) | 540 (83%) | 545 (75%) | 380 (74%) | 305 (75%) | 250 (86%) | 175 (62%) | 175 (79%) |
| EEA | 90 (3%) | 70 (3%) | 40 (6%) | 30 (4%) | 50 (10%) | 25 (6%) | 30 (10%) | 35 (12%) | 15 (6%) |
| IMG | 495 (19%) | 380 (15%) | 70 (11%) | 145 (21%) | 85 (16%) | 80 (19%) | 85 (4%) | 70 (26%) | 35 (15%) |

Numbers rounded as per HESA so may not equal 100%. PMQ= place of medical qualification; UK= United Kingdom; EEA= European economic area; international medical graduates.

**Table 3.** Number of ARCPs by speciality

| **Speciality** | **Satisfactory**<br>**N (%)** | **Insufficient**<br>**evidence**<br>**N (%)** | **Targeted**<br>**training**<br>**N (%)** | **Extended**<br>**training/ left**<br>**programme**<br>**N (%)** | **Total no. of**<br>**ARCPs**<br>**N (%)** |
| --- | --- | --- | --- | --- | --- |
| **General surgery** | 7859 (77.5%) | 1005 (9.9%) | 710 (7.0%) | 562 (5.5%) | 10136 (32%) |
| **Trauma and orthopaedics** | 9098 (82.6%)* | 917 (8.3%) | 638 (5.8%) | 358 (3.3%) | 11011 (35%) |
| **Plastics** | 2091 (85.5%)* | 181 (7.4%) | 89 (3.6%) | 84 (3.4%) | 2445 (8%) |
| **Urology** | 2091 (87.6%)* | 149 (6.2%) | 86 (3.6%) | 62 (2.6%) | 2388 (7%) |
| **Neurosurgery** | 1555 (84.5%)* | 121 (6.6%) | 81 (4.4%) | 83 (4.5%) | 1840 (6%) |
| **ENT** | 1127 (86.5%)* | 81 (6.2%) | 66 (5.1%) | 29 (2.2%) | 1303 (4%) |
| **Maxillofacial** | 812 (81.0%)* | 74 (7.4%) | 47 (4.7%) | 69 (6.9%) | 1002 (3%) |
| **Cardiothoracic** | 782 (83.1%)* | 73 (7.8%) | 37 (3.9%) | 49 (5.2%) | 941 (3%) |
| **Paediatric** | 576 (79.8%) | 60 (8.3%) | 45 (6.2%) | 41 (5.7%) | 722 (2%) |
| **Total** | 25991 (81.8%) | 2661 (8.4%) | 1799 (5.6%) | 1337 (4.2%) | 31788 (100%) |

\* Significant difference in the number of satisfactory outcomes compared to general surgery using $\chi^2$ .

Trauma and orthopaedics had the largest proportion of ARCP outcomes, followed by general surgery (Table 3). There was wide variation in the proportion of non-standard outcomes between specialities (Table 3). General surgery had the lowest proportion of standard outcomes (77.5%), compared to 87.6% in urology. Maxillo-facial surgery had the greatest proportion (6.9%) of extended training/left programme outcomes while ENT had the lowest at 2.2%. All specialities except paediatric surgery had a significantly greater number of satisfactory outcomes than general surgery on Chi square test (Table 3). Figure 1 illustrates the variation in ARCP outcome by speciality. General surgery had the lowest predicted probability of a satisfactory outcome comparing across all specialities and urology had the greatest. General surgery and paediatric surgery had the greatest predicted probability of all other non-standard outcomes.

**Figure 1.**
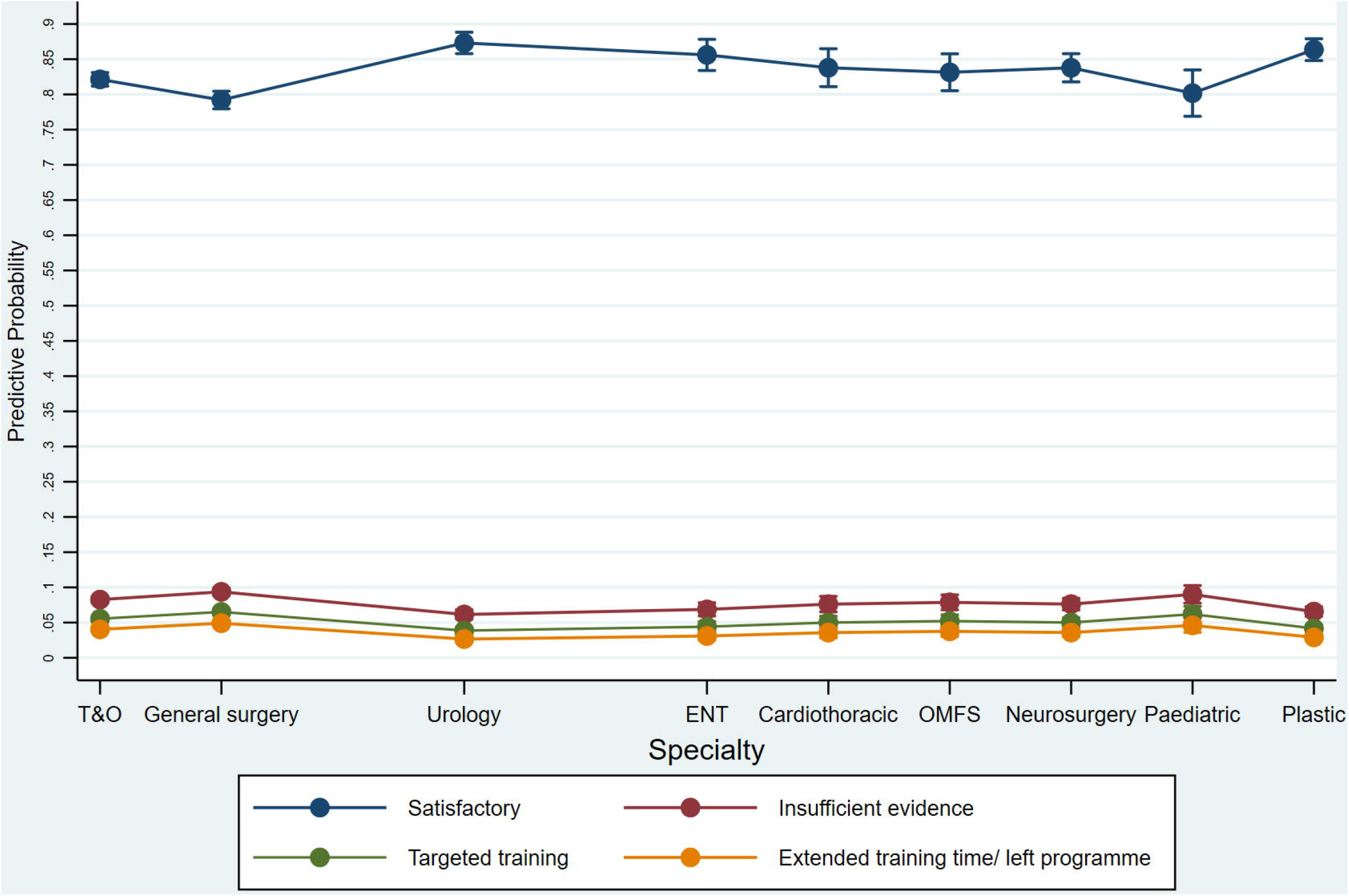
Impact of Specify on ARCP Outcome

On univariable analysis of all specialities combined, factors associated with a non-standard outcome were female sex (OR 1.22 95% CI 1.12-1.33), increasing age at ARCP (1.03 95% CI 1.02-1.04), less than full time training (OR 1.21 95% CI 1.07-1.36) and more recent ARCP date (OR 1.07 95% CI 1.05-1.08) (Table 4). On comparing the subspecialties, plastic surgery (OR 0.81 95% CI 0.70-0.95), urology (OR 0.66 95% CI 0.56-0.78) and ENT (OR 0.75 95% CI 0.61-0.92) trainees were significantly less likely to receive a non-standard ARCP outcome compared to trauma and orthopaedic trainees (Table 4). General surgery trainees were 45% more likely to receive a non-standard ARCP outcome than trauma and orthopaedics (OR 1.45 95% CI 1.32-1.58), and paediatric surgery trainees 28% more likely (OR 1.28 95% CI 1.00-1.63).

**Table 4.** Multi-level ordinal regression showing factors associated with non-standard ARCP outcome.

|  | **Unadjusted OR**<br>**(95% CI)** | **P value** | **Adjusted OR**<br>**(95% CI)** | **P value** |
| --- | --- | --- | --- | --- |
| **Speciality** |  |  |  |  |
| Trauma and orthopaedics | 1 | - | 1 | - |
| General surgery | 1.45 (1.32-1.58) | <0.001 | 1.33 (1.21-1.45) | 0.001 |
| Plastic surgery | 0.81 (0.70-0.95) | 0.010 | 0.70 (0.60-0.82) | <0.001 |
| Urology | 0.66 (0.56-0.78) | <0.001 | 0.64 (0.54-0.75) | <0.001 |
| Neurosurgery | 0.90 (0.75-1.07) | 0.223 | 0.88 (0.74-1.04) | 0.142 |
| ENT | 0.75 (0.61-0.9 2) | 0.005 | 0.76 (0.61-0.93) | 0.009 |
| Maxillofacial | 1.18 (0.95-1.47) | 0.125 | 0.93 (0.75-1.16) | 0.506 |
| Cardiothoracic surgery | 0.96 (0.77-1.21) | 0.753 | 0.88 (0.70-1.10) | 0.268 |
| Paediatric surgery | 1.28 (1.00-1.63) | 0.048 | 1.16 (0.91-1.48) | 0.228 |
| **Female** | 1.22 (1.12-1.33) | <0.001 | 1.11 (1.02-1.22) | 0.020 |
| **Age at ARCP** | 1.03 (1.02-1.04) | <0.001 | 1.04 (1.03-1.05) | <0.001 |
| **PMQ region** |  |  |  |  |
| UK | 1 | - | 1 | - |
| EEA | 0.93 (0.77-1.13) | 0.482 | 0.89 (0.73-1.08) | 0.253 |
| IMG | 0.77 (0.69-0.87) | <0.001 | 0.69 (0.61-0.78) | <0.001 |
| **Less than full time**<br>**training** | 1.21 (1.07-1.36) | 0.002 | 1.11 (0.98-1.26) | 0.093 |
| Yes | 0.50 (0.41-0.62) | <0.001 | 0.51 (0.41-0.63) | <0.001 |
| Missing |  |  |  |  |
| **Year of ARCP** | 1.07 (1.05-1.08) | <0.001 | 1.04 (1.03-1.06) | <0.001 |

Neurosurgery, OMFS and cardiothoracic surgery did not have a significantly higher risk of non-standard outcome compared to trauma and orthopaedics (OR 0.90 95% CI 0.75-1.07, OR 1.18 95% CI 0.95-1.47, OR 0.96 95% CI 0.77-1.21) (Table 4).

After adjustment, women across all surgical specialties were 11% more likely to receive a non-standard ARCP outcome than men (OR 1.11 95% CI 1.02-1.22) (Table 4). For each year increase in age at ARCP, trainees were 4% more likely to receive a non-standard outcome (OR 1.04 95% CI 1.03-1.05). Between 2010 and 2017, there was a 4% increase in the likelihood of non-standard outcome for trainees having an ARCP more recently (OR 1.04 95% CI 1.03-1.06). Compared to UK graduates, international medical graduates were 31% less likely to receive a non-standard outcome (OR 0.69 95% CI 0.61-0.78). There was no significant difference between UK and EEA graduates. Less than full time training did not increase the risk of non-standard outcome across all specialities after adjustment.

After adjusting the data for sex, age, region of primary medical qualification, less than full time training and ARCP year, general surgery trainees were 33% more likely to receive a non-standard ARCP outcome than trauma and orthopaedic trainees (OR 1.33 95% CI 1.21-1.45) (Table 4). Plastic surgery trainees were 30% less likely to receive a non-standard outcome than trauma and orthopaedic trainees after adjustment (OR 0.70 95% CI 0.60-0.82), urology 36% less likely (OR 0.64 95% CI 0.54-0.75) and ENT 24% less likely (OR 0.76 95% CI 0.61-0.93). There was no significant difference for neurosurgery, OMFS, cardiothoracic or paediatric surgery trainees after adjustment.

## Discussion

This is the first study investigating the differences in ARCP outcome between surgical specialties. When combining all surgical specialities, women and older trainees are less likely to receive a standard ARCP outcome. The likelihood of a non-standard outcome has increased throughout time. There is between-specialty variation in ARCP outcomes with General surgery trainees significantly more likely to have a non-standard ARCP outcome compared to other specialities. Across the specialities, 1.3% of trainees are released from training (Outcome 4).

Surgery remains dominated by men and attitudes towards women in surgery have been challenged in recent years. One study reported that 59% of women in surgery had experienced workplace discrimination, there was also a perception that the surgical environment is better equipped to support male trainees[19]. Other studies have found that female trainees have adverse outcomes in surgical training. In a meta-analysis comprising 17,407 surgical trainees across the world, women were found to have a higher pooled attrition than men[20]. A 2019 study found that female general surgery residents in the US were significantly less likely to be granted operative autonomy by faculty than men[6], similar findings were seen amongst New Zealand general surgery trainees[21]. The finding that women across all specialities have adverse outcomes requires investigation at the specialty level.

Between 2010 and 2017, the likelihood of non-standard ARCP outcome has increased. The possible explanations for this include higher standards and criteria of the ARCP panel, changes in training quality and difficulties meeting ARCP requirements. The largest increase in non-standard outcomes was in the insufficient evidence group. This may be due to increased service provision demands leaving less time for portfolio admin and changing curriculum requirements. Previous studies have described a decrease in operative case load for surgical trainees over time[22-24]. A large proportion of UK trainees struggle to meet operative case requirements, with 85% reporting coming in on off days to gain extra experience[25]. The variation in ARCP outcomes over time is likely multi-factorial and beyond the scope of this study.

There are a number of factors which may contribute to the difference in ARCP outcomes between surgical specialties. There are currently differences between requirements for CCT to be awarded between specialties. These inconsistencies have been highlighted in a 2019 paper by Wood et al[26]. For example the academic requirement for CCT in; general surgery requires three peer reviewed publications by the end of training, urology trainees require two, whilst paediatric surgery require four, cardiothoracic trainees are required to deliver six presentations while those in ENT have no specified number. These different thresholds of attainment may therefore contribute to variation seen in the outcomes awarded. Regardless of surgical speciality, there are universal requirements of a day one consultant. The academic and management and leadership responsibilities are likely to be the same across specialities and therefore these should be standardised. The aim of the surgical training programmes is to produce competent day one consultants, this study should prompt a review of these requirements across specialties.

The top performing specialities (urology, plastic surgery and ENT) have a smaller number of trainees which may increase the number of training opportunities and the ease of completing work based assessments. In smaller specialities it is likely that trainees and trainers will have greater training continuity, this has been shown to increase intra-operative entrustment[27 28].

In the United States, general surgery residents have a higher rate of attrition compared to other specialities[29-31]. However, the failure to complete residency in these studies is largely due to voluntary attrition as opposed to unsatisfactory progression. Similarly, in a Canadian study across surgical specialties, general surgery residents were the most likely to consider leaving the programme[32]. The main reason for this was concerns regarding work-life balance. Reassuringly, we have found a low proportion of trainees are asked to leave the programme.

Surgical trainees have been found to have low rates of career satisfaction and higher rates of burnout than compared to trainees in other medical specialties[33]. In a 2019 meta-analysis, general surgery trainees had the highest rate of burnout amongst the surgical subspecialties with a 58% prevalence rate of burnout[34]. Burnout was defined as a long-term stress reaction marked by emotional exhaustion, depersonalization, and reduced personal accomplishment [35]. Orthopaedics and neurosurgery had a slightly lower rate at 55% and 52% respectively[34].

The major strength of this study is that it captures a large contemporary cohort of surgical trainees across specialties. UKMED contains data from reliable sources and avoids the reliance on survey data as studies in this field often do. One limitation is the inability to include vascular surgery within the analysis, due to the small number of ARCP outcomes available. It is not possible to determine the underlying causes for the findings in this study, which are likely to be multifactorial. Further detailed work examining individual ARCP outcomes is required.

## Conclusion

There is a wide variation in the use of non-standard ARCP outcomes across UK surgical specialities with females and older age at ARCP being associated with non-standard outcomes. Furthermore the use of non-standard outcomes is becoming more frequent over time. Possible explanations for this include variation in CCT requirements, training quality or difficulty meeting ARCP standards. The finding that some specialities and groups of trainees have lower rates of standard ARCP outcomes requires further investigation to ensure consistent training quality and assessment regardless of specialty. An in depth study including details of work based assessments, logbook data and supervisor reports per specialty could help to further understand the variation in outcomes.

## Data Availability

The data used in this study is held by UKMED and is available upon request.

## Acknowledgements

Funding will be sought from the University of Nottingham for publication costs associated with open access. There are no conflicts of interest.

## Author contributions

All authors were responsible for editing and drafting the final manuscript. CH performed the statistical analysis.

